# Efficacy and safety of a one-step warming protocol of vitrified blastocyst stage embryos

**DOI:** 10.1101/2025.10.02.25337068

**Authors:** E.R Ebinger, H. Misurac, A.M. Giovannini, V.B. Desai, N. De Rosa, R. Vassena, P. J. Atkinson

**Author notes:** **Correspondence:** Ebinger E.R.

## Abstract

**Objective:** To study the effect of a one-step warming technique on survival and clinical outcomes of vitrified-warmed blastocysts on a consecutive cohort of patients.

**Design:** Retrospective consecutive cohort study.

**Subjects:** This study included 1402 transferred embryos from 989 unique patients that were treated in 3 clinics.

**Exposure:** The exposure group included embryos warmed using a one-step warming protocol of 1M sucrose solution for 1 minute. The control group included embryos warmed using traditional, multi-step warming, where embryos were exposed to 1M sucrose for 1 min, followed by 3 min in 0.5M sucrose, and 10 mins in washing solutions.

**Main Outcome Measures:** The goal of this study was to compare survival, clinical pregnancy (CPR) and ongoing pregnancy (OPR) rates between multi- and one-step warming techniques. Additionally, subgroup analyses by maternal age, embryo morphology, day of vitrification and mode of fertilization were also performed.

**Results:** Survival rates were comparable across all comparisons. Pregnancy rates were comparable between multi-step and one-step groups (CPR: 42.6% vs 44.3%, p=0.78; OPR: 33.2 %vs 37.5%, p=0.21). Pregnancy probabilities between warming techniques were comparable between groups at any age point (32 – CPR: 43.0% vs 47.7%; OPR: 34.5% vs 39.5; p>0.05; 42 – CPR: 24.2% vs 19.3%; OPR: 13.5% vs 13.5%; p>0.05). Good quality embryos (G2) had a lower chance of pregnancy than top quality embryos (G1) overall, but pregnancy rates were similar between groups (G1 – CPR: 52.3% vs 54.6%; OPR: 46.0% vs 48.1%; p>0.05; G2 – CPR: 38.6% vs 40.0%; OPR: 27.8% vs 33%; p>0.05). Similarly, Day 6 embryos were less likely to achieve pregnancy than Day 5 embryos, but pregnancy rates were comparable between groups (D5 – CPR: 44.8% vs 46.5%; OPR: 35.3% vs 40.0%; p>0.05; D6 – CPR: 28.0% vs 31.2%; OPR: 18.3% vs 23.4%; p>0.05). Pregnancy rates between ICSI for male factor infertility and IVF were again comparable between groups (ICSI – CPR: 40.9% vs 38.3%; OPR: 33.7% vs 32.5%; IVF – CPR: 45.0% vs 48.0%; OPR: 37.3% vs 41.9%; p>0.05).

**Conclusion:** One-step embryo warming provides similar survival and pregnancy outcomes compared to classical multi-step warming while decreasing the procedure time by more than 90%.

## INTRODUCTION

Vitrification and warming of embryos are essential tools in assisted reproductive technologies (ART). The introduction of these techniques has improved treatment safety, efficacy and patient outcomes, enabling efficient fertility preservation (1), delayed embryo transfer in cases of elevated progesterone (2) or risk of ovarian hyperstimulation syndrome (OHSS) (3), as well as allowing for uncoupling of ovarian stimulation from embryo transfer in donor cycles (4).

One of the main goals of cryopreservation is preventing ice crystal formation in both intra- and extracellular spaces, to avoid disrupting subcellular structures such as microtubules and organelles, and causing osmotic stress by concentrating solutes in the residual unfrozen solution (5). Vitrification bypasses ice crystal generation through the formation of an amorphous glassy solid by increasing viscosity of a liquid during cooling. Essentially, the solution remains a liquid and retains fluidic properties, but its viscosity is so high that it acts like a solid (6). This process is achieved through high concentrations of cryoprotectants (CPAs) and fast rates of cooling (7). Vitrification utilizes both permeable and non-permeable CPAs. Permeable CPAs, typically small molecules such as ethylene glycol or dimethyl sulfoxide (DMSO), penetrate the plasma membrane and form bonds with water molecules, preventing ice crystal formation. Additionally, permeable CPAs increase the viscosity inside the cells and lower the glass transition temperature. Non-permeating CPAs, such as sucrose and trehalose, remain outside the cell and work by drawing out water and dehydrating the intracellular space. This decreases the available water inside the cell, which minimizes crystallization and reduces the amount of CPAs needed to permeate into the cell. Furthermore, non-permeating CPAs regulate osmotic shifts to prevent osmotic shock and increase the viscosity of vitrification solutions (8). Warming of vitrified embryos consists of the removal of CPAs and rehydration of the cells. This process is equally as delicate as vitrification and requires high warming rates to prevent ice crystallization. Current warming techniques include stepwise rehydration, allowing for the gradual exchange of water and CPAs to prevent osmotic shock and cell rupture (9). Although vitrification and warming have increased the efficacy of ART, the use of high concentrations and prolonged exposure to CPAs remains cytotoxic to oocytes and embryos.

Recent innovations in cryopreservation include shorter vitrification and warming protocols, which minimize both the time embryos spend outside of the incubator and exposure to CPAs. Further, a simplified protocol can increase lab efficiency through shorter procedure times, reduced chance of error and streamlining of training efforts. In particular, there is a focus on the efficacy of a “super-fast”, one-step thaw protocol. For instance, a recent study evaluated a protocol that removes the subsequent dilution steps involved in standard embryo warming and utilizes only one step in 1M sucrose solution, resulting in comparable survival and clinical pregnancy rates, while improving ongoing pregnancy rate and reducing miscarriage rate overall, compared to traditional, multi-step warming techniques (10). Other authors have reported comparable implantation and pregnancy rates between multi-step and one-step warming, while saving embryologists over 14 minutes of time for each warming procedure (11). Other, similar protocols have also been tested, with comparable live birth rates to standard warming (12).

While one-step warming protocols are promising, several unanswered questions remain. For instance, most reports focus on embryos of high morphological grade, from good prognosis patients or oocyte donors. Recently, one-step warming was shown to cause overexpansion of embryos leading to cell lysis (13). One could hypothesize that this cryodamage may become more apparent in embryos of lower morphological grade, especially in the trophectodermal compartment, or those with lower developmental competence such as those developing slower, or from advanced maternal age patients. The aim of this study was to assess the safety and effectiveness of a one-step blastocyst warming technique both in the general and in selected patient populations, such as advanced maternal age, poorer quality embryos and embryos derived from male factor infertility.

## MATERIALS AND METHODS

### Study population

This is a retrospective analysis of a consecutive cohort of 1361 treatment cycles, corresponding to 989 unique patients and 1402 transferred embryos at Adora Fertility labs from August 2023 to April 2025. Up to July 2024, all warming of blastocyst stage embryos in all clinics were performed according to a traditional, multi-step protocol. From July 2024, all embryo warmings were switched to a one-step protocol in all clinics as a matter of clinical practice. All cycles were performed with the patients’ own gametes, and without preimplantation genetic testing. The number of patients and embryos included at each stage of the analysis is presented in **Figure 1**. Women included in this analysis were on average 34.05±4.51 (range: 21-46) years of age at the time of embryo vitrification. Detailed demographic and cycle characteristics can be found in **Table 1 and Figure 2**.

**Figure 1.**
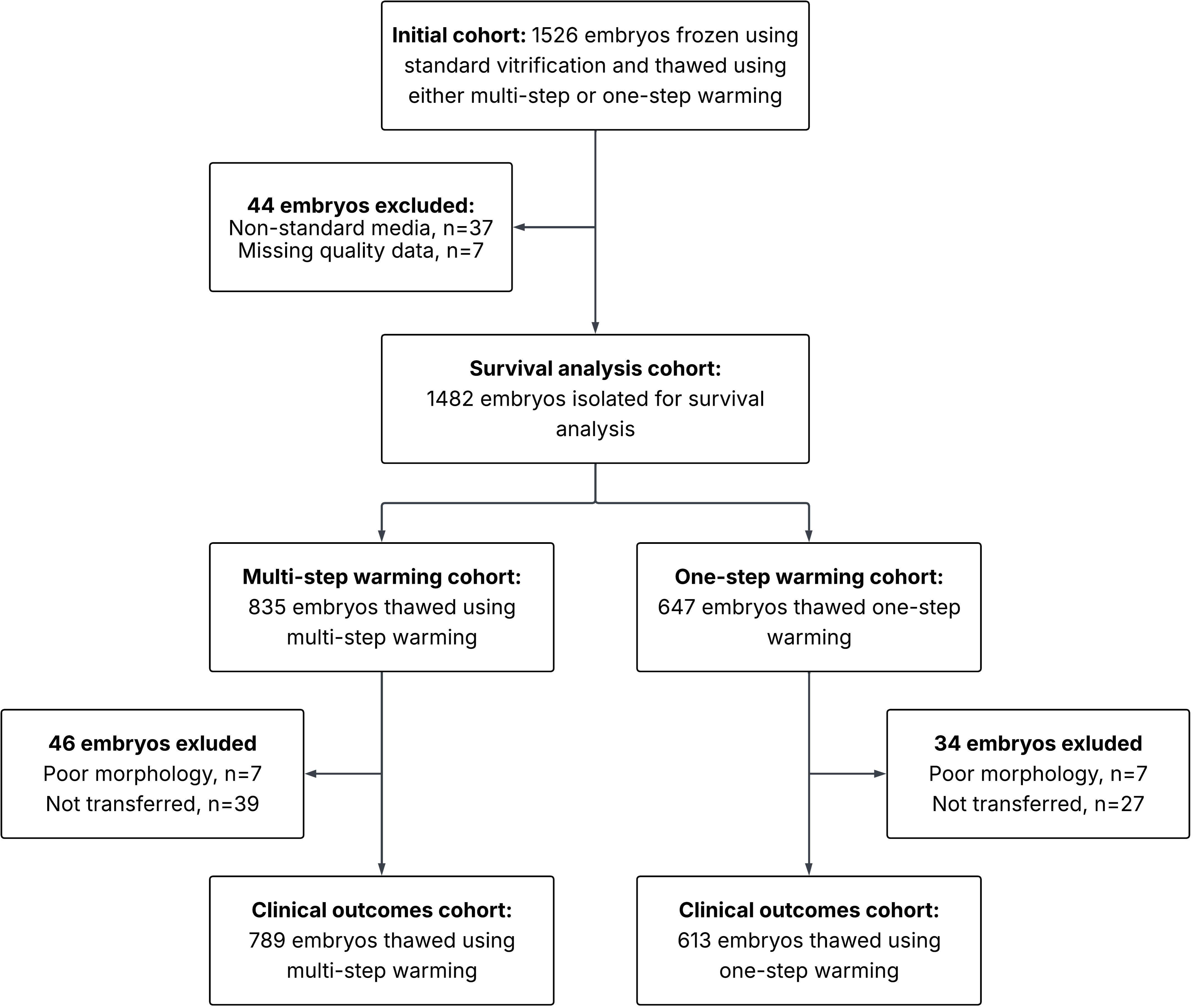
Flowchart of data inclusion and exclusion.

**Figure 2.**
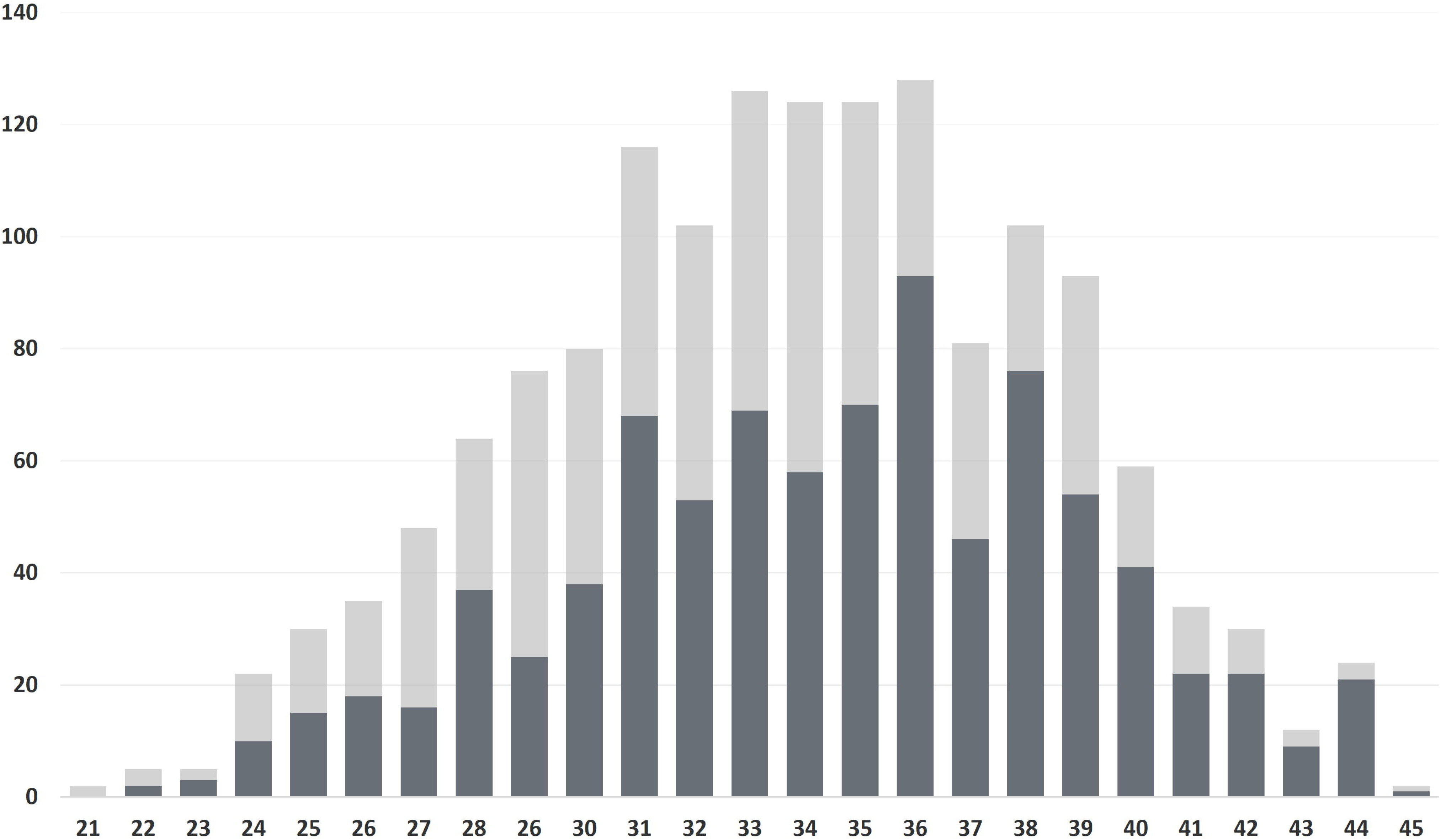
Frequency diagram of age distribution across OSW and MSW groups. X axis: maternal age at vitrification; Y axis: counts. Dark grey bars: MSW; light grey bars: OSW.

**Table 1:** Demographic and cycle characteristics in one-step and multi-step warming groups. Numerical results reported as mean ± standard deviation (range). Categorical variables presented as n (%). BMI = Body Mass Index. ICSI = Intra Cytoplasmic Sperm Injection. FET = Frozen Embryo Transfer.

|  | Multi-step warming | One-step warming |
| --- | --- | --- |
| Patients (n) | 588 | 469 |
| Embryos (n) | 789 | 613 |
| Female age (years) | 34.38 ± 4.56 (22-46) | 33.83 ± 4.5 (21-46) |
| Female BMI (kg/m<sup>2</sup>) | 25.01 ± 4.43 (15.8-40.1) | 24.97 ± 4.36 (16.0-38.0) |
| Male age (years) | 40.67 ± 6.23 (26-69) | 40.52 ± 6.50 (25-75) |
| Sperm motility (a+b;%) | 32.5 ± 27.4 (0-100) | 27.61 ± 24.93 (0-100) |
| Sperm concentration<br>(mill/mL) | 48.05± 42.66 (0.01-250) | 47.9 ± 42.6 (0.01-271) |
| ICSI (% of cases) | 287 (36.40) | 219 (35.70) |
| Natural FET (% of cases) | 467 (59.19) | 361 (58.89) |

### Ovarian stimulation and oocyte retrieval

The most important point to emphasise is that controlled ovarian stimulation regimens and oocyte retrieval procedures did not change over time and therefore the way the embryos were created between the two analysed groups, OSW and MSW, was the same. Patients underwent controlled ovarian hyperstimulation (COH) according to standard and accepted clinical practice. The majority of patients used antagonist protocols. Agonist protocols and flare protocols were used where clinically indicated. The specific type of gonadotropic injection used was according to clinician preference and clinical indication. Treating clinicians at the 3 clinics did not change over time. Oocyte retrievals were completed under heavy sedation according to standardised practice and technique and equipment did not change over time.

### Oocyte fertilization and embryo culture

Sperm samples utilized were either produced fresh or frozen and thawed on day of retrieval. Sperm preparation was performed the same day of oocyte retrieval and was evaluated according to the World Health Organization standards (14).

Following oocyte retrieval, oocytes were inseminated by either conventional insemination (IVF) or intracytoplasmic sperm injection (ICSI). ICSI was only performed if indicated by male factor infertility. Conventional insemination, with 100 x 10^3^ motile sperm/well, occurred 4-6 hours after oocyte retrieval. For ICSI, cumulus cells were removed 2 hours after oocyte retrieval using 80 IU/mL hyaluronidase (Origio SynVitro^TM^ Hyadase, CooperSurgical, Trumbull, USA) and evaluated. Mature (MII) oocytes were then inseminated using ICSI approximately 4 hours after retrieval.

Fertilization was assessed 16 to 18 hours post insemination. Normal fertilization was characterized by the presence of two pronuclei. Zygotes were cultured in culture media (Sage 1-Step, CooperSurgical, Trumbull, USA) overlaid with mineral oil (Sage, CooperSurgical, Trumbull, USA) and cultured in an incubator set to 37°C, 5% CO_2_, 5% O_2_. Blastocyst development was assessed on Day 5 and 6 post insemination. Embryo morphology was assessed based on expansion, appearance and number of cells. Embryos were then allocated to one of three groups. Grade 1 embryos were classified as top quality and Grade 2 embryos were classified as good quality. Equivalence of grades to Gardner scores is presented in **Table 2**.

**Table 2:**
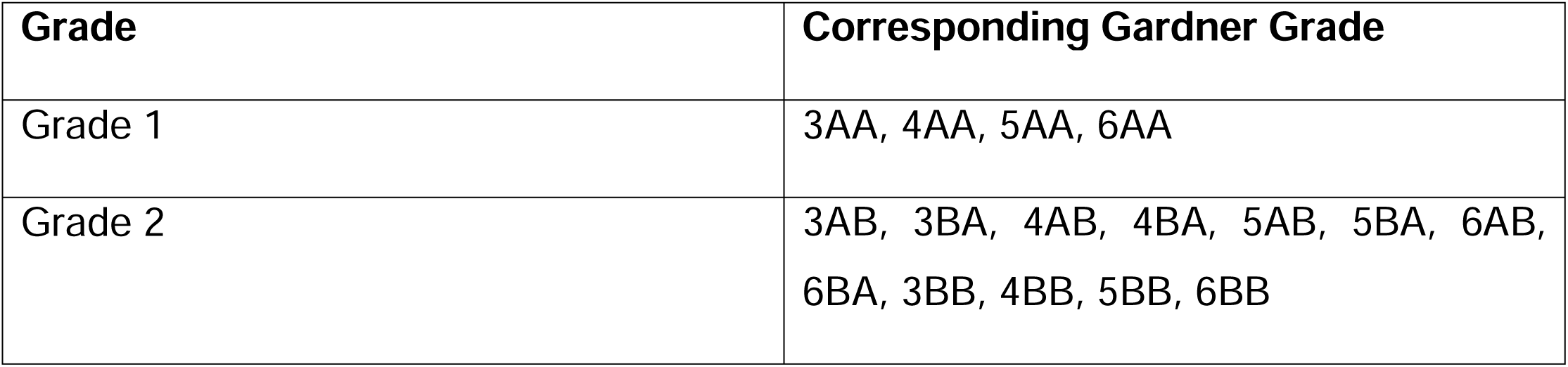
Conversion of embryo morphological grades used in the reported cases and Gardner’s grades.

### Embryo vitrification and warming

#### Embryo vitrification

Embryos were vitrified on D5 or D6 of development. Blastocysts were artificially collapsed by laser pulse at cell-cell junctions of the trophectoderm or by manual collapsing with 175 or 275µm Stripper Tips^TM^ (CooperSurgical, Trumbull, USA) before vitrification. All embryos were vitrified individually and using Vitrifit^TM^ (CooperSurgical, Trumbull, USA). Embryos were placed in solution 1 (10mM HEPES solution) for up to 10 minutes. Embryos were then placed in solution 2 (8% ethylene glycol, 8% DMSO) for 2 minutes and finally exposed to solution 3 (16% ethylene glycol, 16% DMSO and 0.68M trehalose) for 20-30 seconds, before being loaded onto cryopreservation device and plunged into liquid nitrogen. All vitrification procedures were performed at 37^0^C using a blastocyst vitrification kit (Sydney IVF Blastocyst Vitrification Kit, Cook Australia, Brisbane, AUS).

#### Multi-step warming

On the morning of the embryo warming, 1M sucrose was warmed for a minimum of 30 minutes to 37°C, while 0.5M sucrose and MOPs solutions (Sage, CooperSurgical, Trumbull, CT, USA), were warmed to room temperature for a minimum of 30 minutes. Immediately prior to embryo warming, 0.5mL of each solution was aliquoted into separate wells of a 4-well dish (Nunc, Thermo Fisher Scientific, Scoresby, AUS). The cryodevice was removed from liquid nitrogen and plunged into the 1M solution for 1 minute. Subsequently, the embryo was moved to 0.5M sucrose solution for 3 mins. Then, the embryo was passed through two wells of MOPS solution for 5 minutes each. The embryo was then washed through culture media (Sage 1-Step, CooperSurgical, Trumbull, USA) and deposited in 20µL of the same media for recovery in an incubator set to 37°C, 5% CO_2_, 5% O_2_. Transfer of surviving embryos was performed within 1-4 hours of warming.

#### One-step warming

On the morning of the embryo warming, a 1M sucrose solution (Sage Warming Kit, CooperSurgical, Trumbull, USA) was warmed for a minimum of 30 minutes at 37°C. Immediately prior to embryo warming, 0.5mL of warm 1M sucrose solution was aliquoted into a 4-well dish (Nunc, Thermo Fisher Scientific, Scoresby, AUS) and placed on a heating stage. The cryodevice was removed from liquid nitrogen and plunged into the 1M solution for 1 minute. The embryo was gently removed from the device. Subsequently, the embryo was removed from the sucrose solution and washed in culture media (Sage 1-Step, CooperSurgical, Trumbull, USA) and deposited in 20µL of the same media covered with 1.0mL oil pre-equilibrated overnight. Embryos were place in an incubator set to 37°C, 5% CO_2_, 5% O_2_. Embryo transfer of surviving embryos was performed within 1-4 hours of warming.

### Endometrial preparation and embryo transfer (ET)

The most important point to emphasize is that the clinical management of patients and ET technique did not change over time and therefore both cohorts analysed are comparable. Frozen cycle management and ET was performed in line with accepted standard clinical practice.

In ovulatory women, the majority of cycles were natural frozen cycles. In some cycles, a trigger was used if indicated. In anovulatory women, letrozole frozen cycles were used and for some women stimulated frozen cycles were used where clinically indicated. Hormone replacement therapy cycles were used in a minority of cases where clinically required.

ET was performed as per clinical standards using standard embryo placement 1-1.5cm from the uterine fundus under US guidance.

### Statistical analysis

Embryos were categorized into two vitrification groups based on the thawing technique: multi-step warming (MSW) and one-step warming (OSW). Embryos were grouped by grade at freezing, using the classification reported in **Table 2** (Group 1: top quality, Group 2: good quality). Survival after warming was defined as preserved integrity of at least 70% of the embryo 1-2 hours after warming. Biochemical pregnancy was defined as the presence of 50 IU/L bHCG in blood 10 days after ET. Clinical pregnancy was defined as the presence of a beating fetal heart, fetal sac, or otherwise clinically confirmed implantation. And ongoing pregnancy was defined as a beating fetal heart after 12 weeks of gestation.

The primary outcome was ongoing pregnancy, with embryo survival, biochemical pregnancy and clinical pregnancy as secondary outcomes.

All statistical analyses were performed in R (version 4.4.3). To model the relationship between warming protocol and pregnancy outcomes while allowing for non-linear effects of age, a Generalized Additive Models (GAM) was used. The model controlled for additional confounders such as embryo quality and number of transferred blastocysts. To account for the non-independence of multiple embryos from the same patient, a random intercept was included for patient ID. Model estimation was performed using restricted maximum likelihood (REML). Model diagnostics confirmed good fit, and smooth terms were visualized using plot.gam(). The predict() function was used to compute marginal probabilities at fixed ages, and pairwise comparisons of predicted outcomes were conducted to assess the interaction between protocol and pregnancy outcomes at specific maternal ages. A level of 0.05 was selected to determine statistical significance in all cases.

## RESULTS

### One-step warming of embryos and reproductive results

We compared the embryological and reproductive outcomes between embryos allocated to MSW and OSW groups, focusing on survival, biochemical pregnancy, clinical pregnancy and ongoing pregnancy outcomes. This analysis included a total of 1402 transferred embryos from 989 unique patients: 789 embryos from 588 patients in the MSW group and 613 embryos from 468 patients in the OSW group, with 67 patients appearing in both groups. Overall, embryo survival rates were 98.6% for MSW and 98.4%% for OSW groups (p=0.63). The biochemical pregnancy rates were 44.2% for MSW and 46.9% for OSW groups (p=0.55). Clinical pregnancy rates were 41.5% for the MSW and 43.7% for the OSW groups (p=0.74). Finally, ongoing pregnancy rates were 32.2% for the MSW group and 36.1% for the OSW group (p=0.19). Overall, rates for survival, biochemical pregnancy, clinical pregnancy and ongoing pregnancy were comparable between groups when controlling for maternal age, embryo quality and number of transferred blastocysts. Age, as expected, showed a significant, non-linear correlation with pregnancy rates, and the effect of advancing age differed between the two study groups, with a more pronounced effect on the OSW group (edf = 1.8 for MSW and edf = 3.1 for OSW) (**Figure 3**).

**Figure 3.**
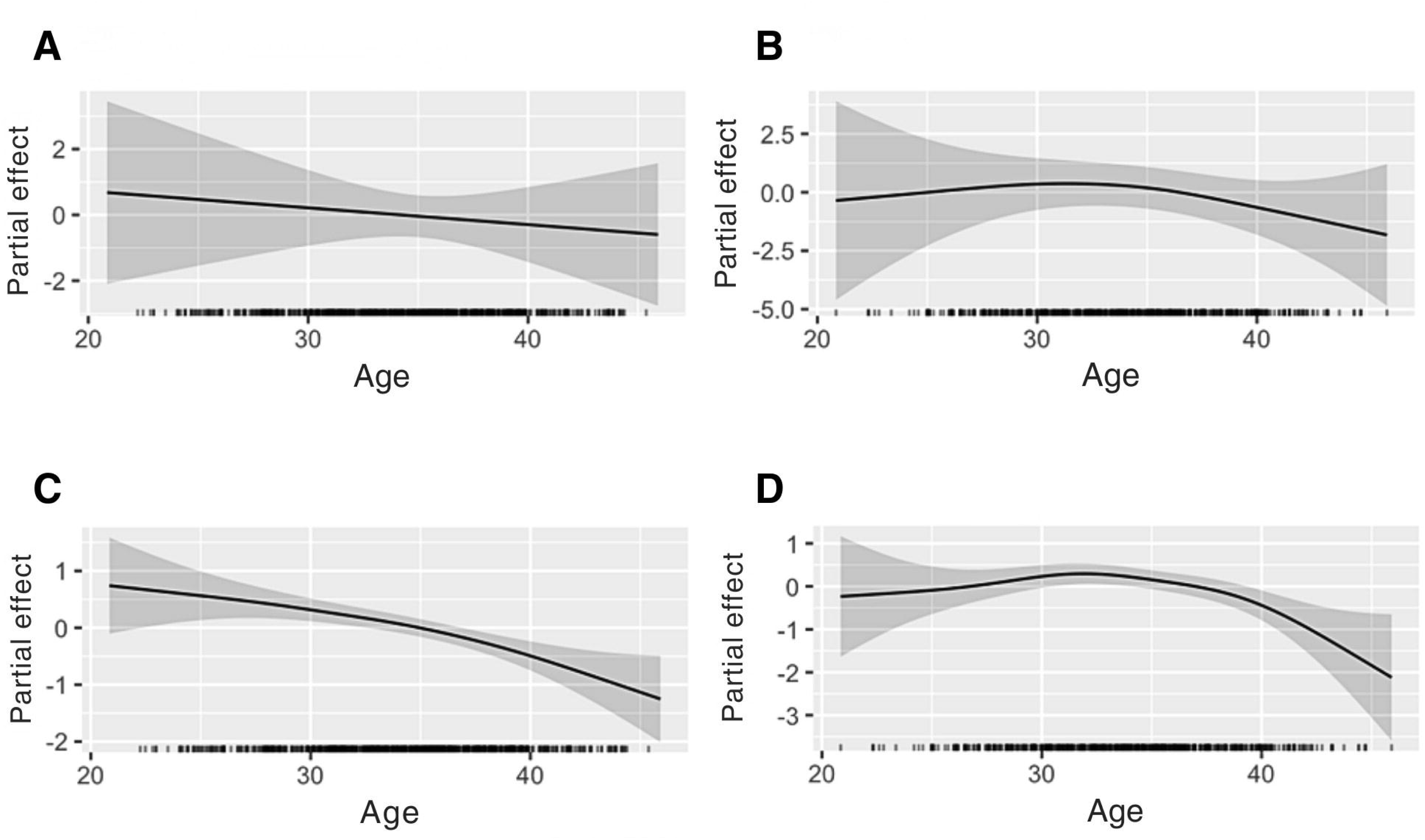
Non-linear effect of age at embryo vitrification on survival and pregnancy outcomes across warming techniques. (A) Effect of age on embryo survival in multi-step warming. (B) Effect of age on embryo survival in one-step warming. (C) Effect of age on pregnancy in multi-step warming. (D) Effect of age on pregnancy in one-step warming.

### One-step warming of embryos from advanced maternal age patients

To evaluate whether maternal age affected embryo survival, biochemical, clinical and ongoing pregnancy chances between warming groups, we predicted the probabilities from the fitted model. Survival rates showed no significant differences across ages and groups (p>0.05 for all comparisons). As expected, we observed a progressive decline in the probability of pregnancy with increasing maternal age in general. For instance, compared to age 32, at age 42 the predicted probabilities for biochemical, clinical and ongoing pregnancy dropped by an average of 28 percentage points in MSW and 33 percentage points in OSW, respectively. However, no significant interaction was observed between groups at any age, indicating that the effect of age on pregnancy probability was comparable between one-step and multi-step warming. For a more detailed view of predicted probabilities at different ages and SART age by warming group, see **Table 3**.

**Table 3:**
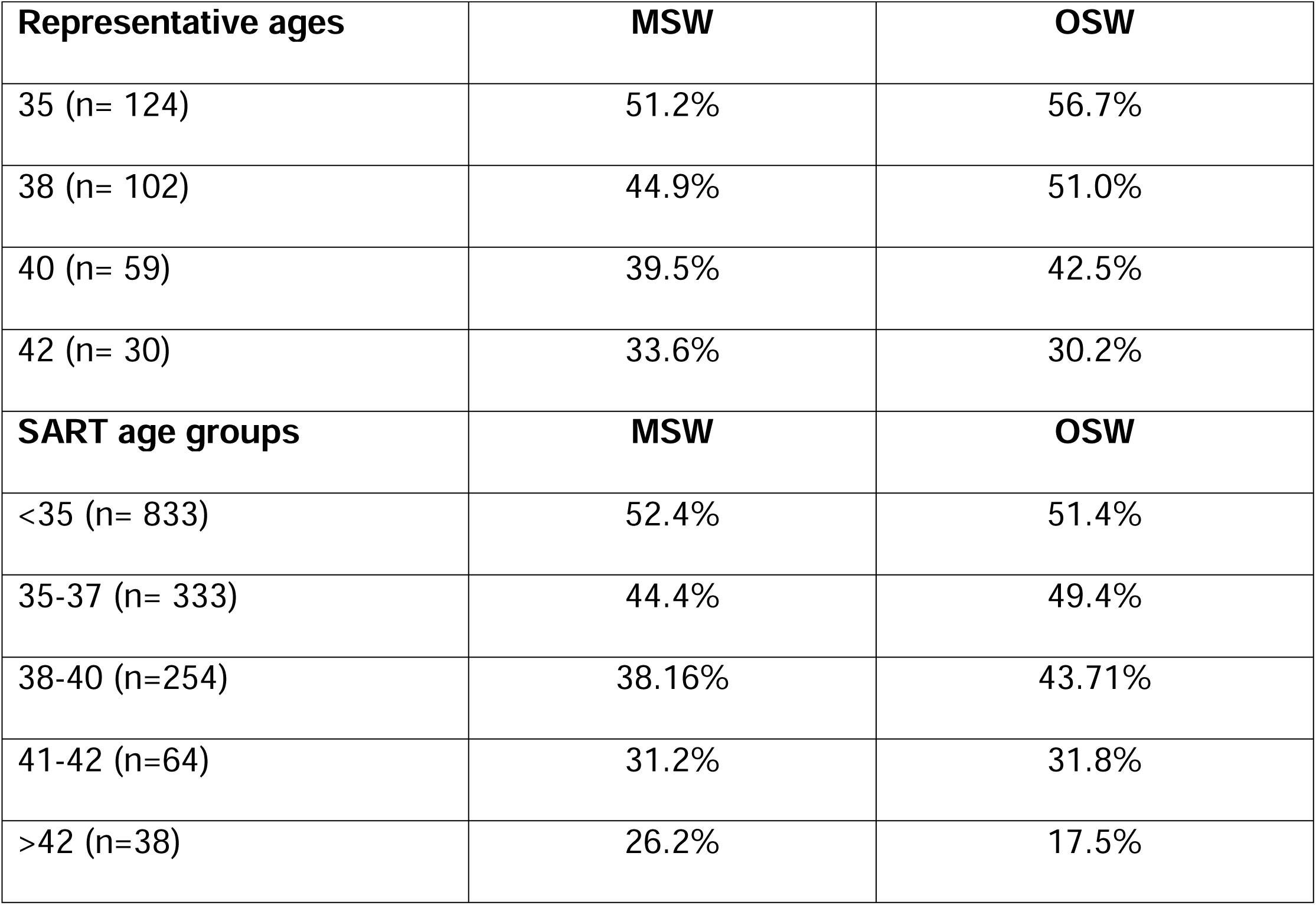
Predicted pregnancy probability at specific ages and in SART age groups by OSW and MSW groups.

### One-step warming and embryos variable morphological grade

Embryos were divided into two groups based on morphological grade. Embryos considered top quality were classified as G1 (MSW: n=235, OSW: n=183), while embryos considered good quality were classified as G2 (MSW: n=554, OSW: n=430). Embryo survival, biochemical pregnancy, clinical pregnancy and ongoing pregnancy rates were analysed by test group and morphological grade. Embryo survival rate for G1 embryos was 98.4% in MSW and 98.9% in OSW groups. Embryo survival rate for G2 embryos was 98.6.% in MSW and 98.2% in OSW groups (p=0.72). Biochemical pregnancy rate for G1 embryos was 56.2% in MSW and 60.1% in OSW groups, while for G2 embryos it was 40.6% in MSW and 42.3% in OSW groups. Clinical pregnancy rate for G1 embryos was 52.3% in MSW and 54.6% in OSW groups. Clinical pregnancy rate for G2 embryos was 38.6% in MSW and 40.0% in OSW groups. Ongoing pregnancy rates were 46.0% in MSW and 48.1% in OSW for G1, and 27.8% in MSW and 33% in OSW for G2 embryos. We observed significantly lower pregnancy rate for G2 embryos across both MSW and OSW groups (45% lower chances of pregnancy p<0.001), but no differences between groups in biochemical (p=0.53), clinical (p=0.80) and ongoing (p=0.83) pregnancy rate.

### One-step warming of slowly developing embryos

Embryos were additionally segregated based on their day of development at vitrification. Embryos vitrified on Day 5 (MSW: n=648, OSW: n=508) and Day 6 of development (MSW: n=93, OSW: n=64) were compared for survival, biochemical, clinical and ongoing pregnancy rates in both MSW and OSW groups. Embryo survival rate for D5 embryos was 98.9% for MSW and 98.8% for OSW groups, while for D6 embryo survival was 98.9% for MSW and 98.5% for OSW groups (p=0.88). Biochemical pregnancy rate for D5 embryos was 47.5% for MSW and 49.4% for OSW groups, while for D6 embryos it was 31.2% for MSW and 35.9% for OSW groups (p=0.7). Clinical pregnancy rates for D5 embryos were 44.8% for MSW and 46.5% for OSW groups and for D6 28.0% for MSW and 31.2% for OSW groups (p=0.12). Ongoing pregnancy rates were 35.3% for MSW and 40.0% for OSW for D5 and 18.3% in MSW and 23.4% in OSW for D6 embryos (p=0.5). Overall, D6 embryos displayed significantly worse reproductive outcomes than D5 embryos when controlling for age and number of transferred blastocysts (50% lower chances of pregnancy, p<0.001), as expected. However, there was no difference between MSW and OSW in biochemical (p=0.75), clinical (p=0.82) and ongoing (p=0.22) pregnancy rates.

### One-step warming of embryos from patients with male factor infertility

Further, reproductive and embryological outcomes were compared based on fertilization method, as ICSI was only performed in case of an indication for male factor. Embryos derived from male factor infertility patients (MSW: n=208, OSW: n=154) and fertilized through ICSI were compared to embryos from non-male factor infertility fertilized through classic IVF (MSW: n=362, OSW: n=277). Embryo survival rates were 98.8% for MSW and 99.1% for OSW groups in male factor cycles, and 99.3% for MSW and 100% in non-male factor cycles, with no differences across groups or indication (p=0.25).

The biochemical pregnancy rates were 43.3% for MSW and 40.3% for OSW groups in male factor cycles, and 48.3% and 51.3%, respectively, in non-male factor cycles (p=0.9). Clinical pregnancy rates were 40.9% for the MSW and 38.3% for the OSW groups in male factor cycles and 45.0% and 48.0% in non-male factor cycles (p=0.7). Ongoing pregnancy rate was 33.7% for MSW and 32.5% for OSW in male factor cycles and 37.3% in MSW and 41.9% in OSW for non-male factor cycles (p=0.4) Overall, rates for survival, biochemical pregnancy, clinical pregnancy and ongoing pregnancy were comparable between groups when controlling for age, embryo quality and number of transferred blastocyst (p=0.3).

## DISCUSSION

“Super-fast”, one-step protocols are poised to streamline operations for clinics, especially considering the continuous increase in frozen embryo transfer cycles worldwide. Traditional, multi-step vitrification and warming, however, are well established techniques with an extremely high survival rate proven across millions of embryos. Responsible innovation mandates that operational improvements are not made at the expense of patient outcomes; it is therefore imperative that these new protocols are not just evaluated in good prognosis patients, but especially in those that stand to lose the most from a decrease in efficacy, i.e. women with few embryos, those with embryos of lower quality or otherwise worse prognosis.

Here, we evaluated 1402 transferred embryos across 3 centers, in one of the largest one-step warming analysis to date and show that, overall, the use of a one-step warming protocol provides comparable results to traditional, multi-step warming protocols. Previous studies have shown that one-step warming provides comparable or improved pregnancy rates to traditional warming techniques (15,16). However, few have addressed the question of the impact on the most fragile embryos. We provide support for the efficacy and safety of one-step warming in subpopulations of embryos that are expected to be more susceptible to cryodamage, such as lower quality, slowly developing embryos or those from patients with male factor or advanced maternal age.

As expected, we identified a trend towards lower pregnancy odds as maternal age increased and embryo quality decreased, however this trend did not affect specifically the embryos warmed with the one-step protocol. A recent study compared pregnancy outcomes between multi- and one-step warming and found comparable implantation, clinical pregnancy, spontaneous abortion and live birth rates after adjusting for maternal age, among other variables (17). Our work extends and improves previous reports: importantly, the effect of maternal age on reproductive outcomes is not linear; for instance, the effect of age between 33 and 35 years old is less pronounced than the effect between 38 and 40. Age should therefore be modelled considering this non-linearity. We found that the effect of the one-step warming procedure is also non-linear, something that has not been previously described. As a result, we employed GAMs that allow for the analysis of multiple non-linear effects, unlike other published studies, and provide a more accurate assessment of the warming protocol effect on reproductive outcomes.

Further we found that embryos of poorer quality have reduced rates of pregnancy, however, this was observed across both groups. When comparing multi-step warming to one-step warming the results for survival, biochemical, clinical and ongoing pregnancy were comparable. Our results support and extend those of a study comparing the rates of embryo survival, clinical pregnancy, negative pregnancy, biochemical/ectopic pregnancy, and spontaneous abortion between one-step and multi-step warming techniques which also found no differences in outcomes when comparing good-with poor-quality embryos (18).

Moreover, we show that one-step warming remains effective on slower developing embryos that have been vitrified on D6. Our results align with those of a very recent study, where multi-step and one-step warming procedures were stratified based on development day, and found comparable clinical pregnancy rates (19).

Finally, we focused on embryos derived from poor quality sperm, where ICSI was indicated. A mounting body of evidence indicates that the contribution of the sperm to the developmental potential of the embryo is substantial (20,21) and may affect the embryo’s response to vitrification and warming protocols. We found comparable rates for survival, biochemical, ongoing and clinical pregnancy between multi- and one-step warming protocols. This is the first time that the contribution of the sperm is taken into account in the evaluation of one-step warming and further reassures in its safety.

Although the results from the present study are promising, we acknowledge some limitation in our study. First, as in all retrospective studies, some relevant variables may be unaccounted for in the controlled analysis, and this may affect the observed outcomes. Additionally, live birth results are still pending. Finally, the data presented here were collected in one group of associated clinics. This allowed for the inclusion of a high number of cycles, while controlling for variation in protocols and practices, although it may limit the generalizability of results.

Given the reliance on retrospective data in this area of innovation, there is a clear need for prospective studies to further elucidate the possible differences. Additionally, future studies should continue to consider data related to patient diagnoses, especially as they pertain to lower quality embryos.

## CONCLUSION

In conclusion, our work supports the potential of “super-fast”, one-step blastocyst warming techniques. One-step blastocyst warming leads to comparable survival, biochemical and clinical pregnancy rate, while lessening procedure time, CPA exposure, and room air exposure, and strengthening the chain of custody and lowering the chance of error.

## DECLARATION

### Data availability

The data underlying this article are available upon reasonable request to the corresponding author.

## Acknowledgements

We would like to thank Filippo Zambelli of TRT Consulting for statistical support.

## CRediT Authorship Contribution Statement

**Emma R. Ebinger:** Conceptualization**, Supervision,** Writing – review & editing, Data validation. **Helena Misurac:** Data curation. **Alexa M. Giovannini:** Writing – original draft, review & editing, data visualization. **Vrunda B. Desai**: Writing – review & editing, Resources. **Nadia De Rosa:** Writing – review & editing. **Rita Vassena:** Writing – review & editing, Methodology, Formal analysis, Conceptualization. **Paul J. Atkinson:** Writing – review & editing.

